# Effects of Postnatal Glucocorticoids on Brain Structure in Preterm Infants, A Scoping Review

**DOI:** 10.1101/2022.11.09.22282133

**Authors:** Isabella Robles, Margarita Alethea Eidsness, Katherine E. Travis, Heidi M Feldman, Sarah E. Dubner

## Abstract

Glucocorticoids (GC) are used in neonatal intensive care units to prevent or reduce the severity of chronic lung disease in preterm infants and have been implicated in impaired neurodevelopment. Our objective was to identify what is known about the effects of postnatal GC treatment in human preterm infants on structural brain development and to identify gaps in the literature. Following Arksey and O’Malley’s scoping review methodological framework, we searched scientific literature databases for original research on human preterm infants, postnatal GCs, and brain structure. 11 studies assessed the effects of GCs on structural brain outcomes. 56 studies reported brain injury, but not structure. Dexamethasone was consistently associated with decreased total and regional brain volumes, including cerebellar volumes. Hydrocortisone was often, but not always associated with absence of brain volume differences. No studies examined the impact of inhaled GC on brain structure. Additional research on the effects of neonatal GCs after preterm birth on a variety of structural brain measures is required for understanding contributions to neurodevelopment and informing practice guidelines.

## 1.0 INTRODUCTION

Glucocorticoids (GC) are a class of steroid hormones that may be produced endogenously by the adrenal gland or provided exogenously as an anti-inflammatory or immunosuppressant medication. Exogenous GC are highly effective as anti-inflammatory medications, but are associated with multiple complications, especially after prolonged use and affect brain development via intracellular glucocorticoid and mineralocorticoid receptors. Infants born preterm (PT), particularly those born more than 2 months before their due date, labelled very and extremely preterm, often require support for basic bodily functions, such as mechanical ventilation for respiratory illness. These PT children are at high risk for inflammatory complications during their neonatal hospitalization, including a severe inflammatory chronic lung disease, called bronchopulmonary dysplasia (BPD). The use of exogenous GC for infants born preterm (PT) in the neonatal intensive care unit has been a controversial topic, much debated in recent decades. GCs are commonly administered to PT infants to reduce the incidence and severity of BPD, which is a major risk factor for mortality and neurodevelopmental disability in children born PT (Gallini et al., 2021; Patra et al., 2017). The overall aim of this scoping review is to review the existing literature in human infants to determine what is known about the impact of exogenous GC, administered in the neonatal intensive care unit in relation to early brain structural development.

Multiple systematic reviews have considered the effects of and made recommendations for GC administration in preterm infants (Chang, 2014; Doyle et al., 2021a, 2021a; Noguchi, 2014; Onland et al., 2017; Shah et al., 2017). Several reviews have looked at imaging studies and GC as part of an overall review of steroid effects on the neonatal brain, the most recent of which are nearly 10 years old (Baud, 2004; Chang, 2014; Favrais et al., 2014; Noguchi, 2014; Rademaker and de Vries, 2009). The reviews to date have not fully considered the effects of postnatal GCs on developing brain structures, even though alterations in brain structure may be major contributors to adverse neurodevelopmental outcomes (Volpe, 2009). It remains critically important to understand any impacts because changes may relate to later neurodevelopmental outcomes. Indeed, structural brain findings may provide a proximal indicator, available to clinicians while PT children are still hospitalized, of the neurobiological effects of GCs, informing clinical research and practice. Given the increase in neuroimaging availability over the past decade, we sought to review the current literature on GC administration in PT infants and structural brain metrics. Because of the variety of GC types, doses, timing, duration, and the diversity of brain outcomes reported, a scoping review of the literature was most appropriate. The primary objective of this scoping review is to identify what is known about the effects of GC treatment on brain structural development in PT human infants with the goal of identifying gaps in the literature and potential intermediate biomarker candidates for neurodevelopmental outcomes.

## 2.0 METHODS

We followed the Arksey and O’Malley methodological framework for scoping reviews to ensure an orderly approach to mapping the existing evidence on what is known broadly about this topic and to identify gaps in the literature (Arksey and O’Malley, 2005). The stages include: (1) identifying the research question; (2) identifying relevant studies; (3) selecting the studies; (4) charting the data; and (5) collating, summarizing, and reporting the results.

A search query was designed to yield articles that included 3 key topics: postnatal steroids, prematurity, and brain structure. We used a combination of MeSH terms and text word search of titles and abstracts. Our full search terms are shown in the **Supplement**. Using this strategy, we collected articles from ClinicalTrials.gov and PubMed published between January 1, 1990 and September 16, 2021. We also included references from recent Cochrane systematic reviews on postnatal glucocorticoid administration for preterm infants (Doyle et al., 2021a, 2021b; Onland et al., 2017).

The inclusion and exclusion criteria are shown in **Table 1**. We defined a structural brain outcome as description or measurement of one or more anatomical features, assessed using one or more neuroimaging modalities. For example, MRI was commonly used to report on overall and regional brain volumes or image slice area.

**Table 1.** Inclusion and exclusion criteria for title and abstract review

| **Inclusion Criteria** | **Exclusion Criteria** |
| --- | --- |
| 1. Papers from 1990 and later<br>2. English language<br>3. Published and peer-reviewed<br>4. Contains empirical data<br>5. Human subjects<br>6. Infants born before 37 weeks gestation<br>7. Postnatal exogenous GC exposure<br>8. GCs administered systemically - enteral, intravenous, intramuscular, inhaled, nebulized, intranasally administered, sublingual, or subcutaneous<br>9. GCs administered to infants during birth hospitalization<br>10. At least one structural brain outcome measured after GC administration | 1. Conference abstracts, case studies/reports if less than 10 subjects, dissertations; review papers; preprints; theoretical papers<br>2. Endogenous GC exposure or measurement<br>3. GCs administered antenatally only<br>4. No structural brain outcomes reported<br>5. Functional brain imaging studies only<br>6. Animal study with no human participants<br>7. Topical GC application only<br>8. GCs after birth hospitalization discharge |

Studies that reported on structural brain outcomes after postnatal GC administration to preterm children were our primary interest in this review. We differentiated the anatomical descriptions and measurements from general descriptions, such as intraventricular, brain parenchymal, and/or cerebellar hemorrhages or injuries, which may or may not lead to variation in brain anatomy. These findings are frequently used to characterize preterm study populations and are not necessarily treated as an outcome of interest. We defined these outcomes as “brain injury”, reviewed them as a set, and, to provide a comprehensive view of the literature, included these references in the **Supplement**. Studies that included postnatal GC administration as a covariate or as a possible risk factor moderating relations between preterm birth and brain structure, but did not include information on GC type, dose, or administration were reviewed and also included in the **Supplement**. We excluded Near Infrared Spectroscopy (NIRS) and Doppler imaging modalities because they are considered functional modalities.

References were imported into Covidence (“Covidence systematic review software,” n.d.) and were screened by 3 independent reviewers (IR, AE, SD) at each step: title and abstract screening, full-text review, and data extraction. For title and abstract screening, references were excluded only if it obviously met exclusion criteria, such as being an animal study. For full-text review, articles were included if any structural brain outcome was reported, regardless of it being a primary or secondary outcome. The senior author (SD) reviewed and resolved conflicts when consensus through discussion was not reached.

We extracted the following data from relevant studies: study design, aim of study, trial name, cohort size, steroid administration (type, dose, route, duration), imaging modality, primary and secondary outcomes, and brain outcome. For studies in the supplement, the reported outcomes were not structural brain outcomes. For these studies, we extracted the relevant brain data that were reported as a characteristic of the study population following steroid exposure.

## 3.0 RESULTS

Using the criteria described in the methods, we identified 11 papers for inclusion in this review. A consort diagram is shown in the **Figure. Table 2** includes the studies that specifically examined structural brain outcomes as a result of GC exposure. An additional 62 studies were either glucocorticoid intervention trials that reported brain injury outcome (e.g. intraventricular hemorrhage, IVH) as a characteristic of the study population (n= 50), or were observational studies of brain structural outcomes that examined postnatal GC as a potential contributing or risk factor (n=12) and provided little or no information on glucocorticoid type or administration. These studies are summarized in the **Supplement**.

**FIGURE.**
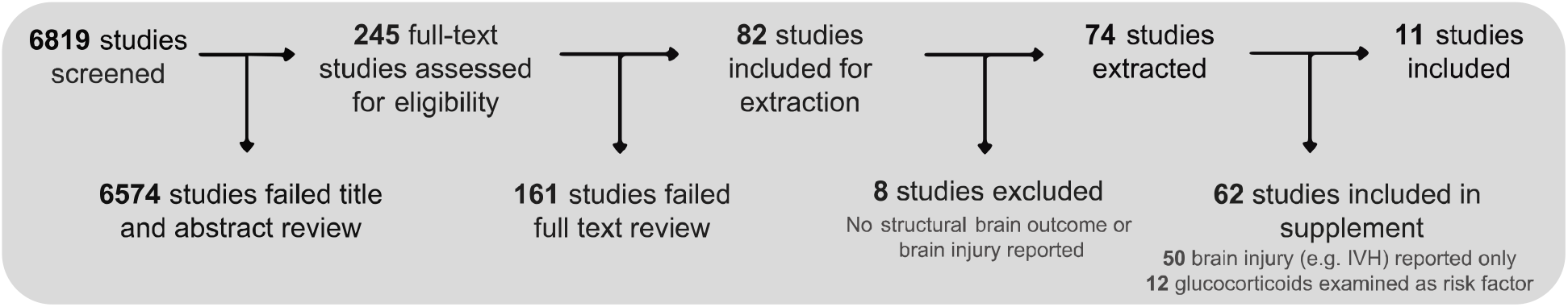
Consort diagram of studies screened and extracted.

**Table 2.** Studies of glucocorticoids and brain structure in preterm born infants

| Steroid | Author Year | Study Type | Number of Participants | Administration Details | Age at Imaging | Structural Brain Outcomes | Effect |
| --- | --- | --- | --- | --- | --- | --- | --- |
| DEXAMETHASONE | Cheong 2014<sup>1</sup> | Cohort Study | GC: 55<br>No GC: 93 | 7.9 ± 4.0 mg/kg for median duration 27d 17-39 either parenterally or orally. | 18 years | Total and regional volumes, hippocampal volume | Reduced brain volume |
|  | Murphy 2001<sup>2</sup> | Cohort Study | GC: 7<br>No GC: 11<br>Term Infants: 14 | Median dose of 0.25 (0.19-0.90) mg/kg/d. With a mean duration of 28d (6-57). Age at first dose: 5 of 7 started in the first 14d of life. | 38-41 weeks post conceptional age | Cortical gray matter, cerebral tissue | Reduced brain volume |
|  | Parikh 2007<sup>3</sup> | Cohort Study | GC: 11<br>No GC: 30 | Mean cumulative dose of 2.8 (1.2-5.9) mg/kg. With a mean duration of 6.8d (2-14). Orally or injected. Age at first dose: 28d. | GC: mean: 41.7 weeks post menstrual age; SD: 3.8 weeks<br>No GC: mean: 38.7 weeks post menstrual age; SD: 2.4 weeks | Total brain, cerebellum | Reduced brain volume |
| HYDROCORTISONE | Alison 2020<sup>4</sup> | RCT | GC: 148<br>No GC: 147 | 1mg/kg/d divided into 2 doses for 7d. 0.5 mg/kg/d for 3d, intravenously. | "Term Equivalent Age" | Kidokoro scoring of cerebral white matter, cortical and deep gray matter, and cerebellar abnormalities; lateral ventricular diameter, biparietal width, interhemispheric distance, deep gray matter area, transcerebellar diameter, vermian height and diameter | No association |
|  | Benders 2009<sup>5</sup> | Cohort Study | GC: 19<br>No GC: 19 | 5 mg/kg/d divided into 4 doses for 1 week, followed by a tapering course of 3, 2, and 1 dose(s) every 5d. Total dose: 3.75, 2.5, and 1.25 mg/kg/d.<br>Age at first dose: 14d (7-44) | GC: mean: 41.1 weeks post menstrual age; SD: 0.8 weeks<br>No GC: mean: 40.8 weeks post menstrual age; SD: 1.0 weeks | Total cerebral tissue and intracranial volume | No association |
|  | Kersbergen 2013<sup>6</sup> | Cohort Study | GC: 73<br>No GC: 73 | 5 mg/kg/d divided into 4 doses for 1 week and a subsequent tapering course every 5d. Total cumulative dose: 72.5 mg/kg over 22 days.<br>Age at first dose: >7d postnatal age | GC: mean: 41.1 weeks post menstrual age; SD: 0.8 weeks<br>No GC: mean: 41.1 weeks post menstrual age; SD: 0.8 weeks | Total brain tissue and cerebellar volume | No association |
|  | Lodygensky 2005<sup>7</sup> | Cohort Study | GC: 23<br>No GC: 35<br>Healthy Term: 21 | 5 mg/kg/d, 4 doses for the first week, followed by a taper (not reported). Median age at first dose: 18d (4-43). | Mean: 8 years 7 months, SD: 8.6 months | Gray matter, white matter, hippocampal volumes | No association |
|  | Parikh 2013<sup>8</sup> | RCT | GC: 31<br>No GC: 33 | 3mg/kg/d q12hrs for 4d, 2mg/kg/d for 2 days, then 1mg/kg/d for 1 day. Total cumulative dose of 17 mg/kg, intravenously. Age at first dose: not reported. At least 7d. | 38 weeks post menstrual age | Total brain tissue volume | No association |
|  | Rademaker 2007<sup>9</sup> | Cohort Study | GC: 62<br>No GC: 164 | 5 mg/kg/d, divided into 4 doses for 1 week. Followed by a tapering course of 3, 2, and 1 dose(s) each for 5D (total of 3.75, 2.5, and 1.25 mg/kg/d, respectively). Median duration: 27.5d (IQR, 12d).<br>Age at first dose: >1 week of postnatal age. | Median 8.1 years | MRI lesions at school age classified as normal, mildly abnormal, severely abnormal; corpus callosum area on a midsagittal T2 | No association |
|  | Rousseau 2021 | Cohort Study | GC 20<br>No GC: 40 | for refractory arterial hypotension: 6-12 | GC: mean: 40 weeks post menstrual | basal ganglia and thalamus area | No association |
|  |  |  |  | mg/kg/d.<br>for life-threatening respiratory insufficiency/prolonged mechanical ventilation beyond 21st day of life: 5 mg/kg/d.<br>median duration of treatment was 8d. Median age of treatment was 12d old for hemodynamic indications (n = 7) and 27d old for respiratory indications (n = 16). | age (37 weeks 0 days – 48 weeks 6 days)<br>No GC: mean: 40 weeks post menstrual age (37 weeks 4 days – 45 weeks 5 days) |  |  |
| HC or DEX | Tam 2011<sup>11</sup> | Cohort Study | Site 1: 57 subjects (92 scans)<br>Site 2: 115 subjects (206 scans) | HC: 1-3 mg/kg/d followed by tapering daily doses.<br>DEX: 0.15mg/kg/d followed by tapering daily doses for BPD or 0.15mg/kg/dose for up to 3 doses prior to extubation for airway edema. | Site 1: first scan mean 31.7, SD 1.6 weeks post menstrual age. Second scan mean: 35.7, SD 2.2 weeks post menstrual age.<br>Site 2: first scan mean 32.3, SD 3.3 weeks post menstrual age. Second scan mean 40.5, SD 2.9 weeks post menstrual age | Cerebellar volumes | Impaired cerebellar growth |

Abbreviations: HC Hydrocortisone, DEX dexamethasone, RCT randomized controlled trial, BPD bronchopulmonary dysplasia
<sup>1</sup>Cheong JL, Burnett AC, Lee KJ, Roberts G, Thompson DK, Wood SJ, Connelly A, Anderson PJ, Doyle LW, 2014. Association between postnatal dexamethasone for treatment of bronchopulmonary dysplasia and brain volumes at adolescence in infants born very preterm. *J Pediatr* 164, 737-743.e1.
<sup>2</sup>Murphy BP, Inder TE, Huppi PS, Warfield S, Zientara GP, Kikinis R, Jolesz FA, Volpe JJ, 2001. Impaired cerebral cortical gray matter growth after treatment with dexamethasone for neonatal chronic lung disease. *Pediatrics* 107, 217–21. https://doi.org/10.1542/peds.107.2.217
<sup>3</sup>Parikh NA, Lasky RE, Kennedy KA, Moya FR, Hochhauser L, Romo S, Tyson JE, 2007. Postnatal dexamethasone therapy and cerebral tissue volumes in extremely low birth weight infants. *Pediatrics* 119, 265–72. https://doi.org/10.1542/peds.2006-1354

### 3.1 DEXAMETHASONE

Three studies reported on the association between dexamethasone exposure and a range of brain structural outcomes across a range of ages at the time of imaging. An early 3-D quantitative volumetric MRI study that imaged the participants at 38-41 weeks post conceptual age compared infants treated with dexamethasone to infants who were not exposed to GC and to term infants (Murphy BP et al., 2001). The dexamethasone-treated group was born at an earlier gestational age, had a longer length of stay, had higher clinical risk index for babies (CRIB) score in the first 12 hours of life. Cerebral cortical gray matter volume was reduced in the dexamethasone-exposed group compared with the non-exposed group, after adjusting for gestational age at birth and CRIB score (mean difference 65.3, 95% confidence interval: 4.3 to 114.58). There were no significant cerebral cortical gray matter volume differences between non-exposed preterm infants and control. Total cerebral tissue volume was reduced by 22% in the dexamethasone treated versus non-treated preterm groups. After adjustment for covariates, the volume remained lower, but was no longer statistically significant. No differences were observed in the subcortical gray matter volumes (basal ganglia and thalami). Myelinated white matter volume differed between preterm and full term groups, but was not different between dexamethasone exposed and non-exposed preterm infants. Unmyelinated white matter did not differ between groups.

Parikh and colleagues (Parikh NA et al., 2007) examined a later sample with lower overall dexamethasone doses, adjusting for multiple potential confounders (PMA at scan, dexamethasone treatment, birthweight, presence of BPD). They obtained coronal T2 weighted MRI at a mean post menstrual age of 39.4 weeks. The authors reported smaller total brain volumes (9.5%), cerebellar volumes (19.7%) and subcortical gray matter volumes (20.6%) in the dexamethasone group. Total gray matter volume was also lower in the dexamethasone group but did not meet statistical significance. There was no dose response relationship between dexamethasone dose and total brain volume and cortical tissue volume.

A third study obtained T1 weighted MRI at age 18 years in a cohort of individuals born preterm who had received NICU administered dexamethasone (Cheong et al., 2014). In this cohort, GC were given at the discretion of the treating clinicians after the first week of life. The dexamethasone group had a lower gestational age and birthweight and higher rates of BPD and cerebral palsy than those who did not receive postnatal dexamethasone. Overall they found a 3.6% (95% CI -7.0%, -0.3%) smaller total brain tissue volume at age 18, that was not significantly different between groups after adjustment for covariates. Volumes of total and most regional areas of cortical white matter, thalamus, and basal ganglia nuclei were smaller in the dexamethasone group compared with the no-dexamethasone group after adjustment for covariates. Unlike the studies reviewed here which looked at scans from younger ages, there was no statistically significant difference in cortical gray matter, hippocampus, amygdala, and cerebellar volumes. Within white matter, on the other hand, the reduction in brain parcel volumes in the dexamethasone group was found in most regions except the medial temporal region.

### 3.2 HYDROCORTISONE

Eight studies directly assessed the association between hydrocortisone and brain structure.

Two studies reported on overlapping samples of PT infants with 3D volumetric MRI at term equivalent age. Benders and colleagues performed a prospective study comparing infants who received a two-week course of hydrocortisone at 1 week of life or later with age, gender, and respiratory status matched controls from a second institution using the same MRI scanner and the same imaging protocol (Benders et al., 2009). Adjusted analyses showed no differences between groups for individual or summed volumes for the intracranial cavity, CSF, cortical gray matter, subcortical gray matter, unmyelinated white matter, and myelinated white matter. In the second investigation of an expanded cohort (Kersbergen et al., 2013) there was no difference in total brain tissue volume or cerebellar volume between treated and untreated controls after adjustment for covariates. This was the same whether hydrocortisone was treated as a continuous variable or high vs low dose.

In a pilot randomized controlled trial, Parikh and colleagues obtained axial PD/T2 volumetric MRI scans at 38 weeks post menstrual age in infants randomized to hydrocortisone or placebo with the specific objective of examining a structural brain outcome (Parikh et al., 2013). Total brain tissue volume was the primary outcome.

Secondary brain outcomes included individual tissue volumes of cortical gray matter, cerebral white matter, cerebrospinal fluid, subcortical gray matter and substructure volumes of included nuclei, cerebellum, hippocampi, amygdalae, corpus callosum, and brain stem. The study was powered to detect a 2-week growth difference in brain size. In bivariate and adjusted analyses, total tissue volume was not different between the groups. Secondary brain outcomes of regional volumes were not different between the groups.

Alison et al. (Alison M et al., 2020) conducted a large predefined secondary analysis of T1, T2, T2 GE MRI at term equivalent age of patients enrolled in the PREMILOC trial of early hydrocortisone. Kidokoro scores were used to evaluate cerebral WM, cortical and deep GM and cerebellar abnormalities (Kidokoro et al., 2013). The primary endpoint was the occurrence of cerebral white matter abnormalities. There were no differences in the cumulative distribution of white matter scores between the treatment and control groups. Fewer infants born at 24-25 weeks in the hydrocortisone group developed dilated lateral ventricles compared with the placebo group. There were no differences in any of the cortical and basal ganglia gray matter, nor cerebellar injury between the groups. There was a statistically significant association between overall brain injury score cumulative distribution between the groups, however hydrocortisone was no longer significantly associated with white matter damage or overall moderate to severe brain damage after adjustment for risk factors and gestational age.

Rousseau (Rousseau et al., 2021) obtained axial T2 MRI and aimed to explore postnatal brain growth in extremely preterm born infants requiring postnatal steroids. They examined brain tissue area at term equivalent age and head circumference growth by 12 months of age. This report included children receiving hydrocortisone for any indication (e.g., refractory hypotension, prolonged respiratory insufficiency).

Semiautomatic tissue segmentation was performed and areas of each tissue were measured on an axial slice (not a 3D volumetric measurement). The study was powered to see an 8% reduction of brain volumes at term equivalent age. Despite matching, the hydrocortisone group was lower weight at birth and had more medical complications.

Adjusting for postmenstrual age at MRI scan, the hydrocortisone treated group had a significant reduction in the intracranial cavity area and the basal ganglia and thalamus area at term equivalent age. However, there was no dose-dependent relationship between the amount of hydrocortisone and adjusted basal ganglia and thalamus values. Multiple regression showed that duration of mechanical ventilation was the only significant independent variable associated with BGT (not with hydrocortisone treatment).

Lodygensky et. al obtained 3D quantitative volumetric MRI in 8 year old children who had been born preterm or full term (Lodygensky et al., 2005). A portion of the PT children had received hydrocortisone due to respiratory illness severity. No differences were seen between PT children who did or did not receive neonatal hydrocortisone in total intracranial volume, gray matter volume (cerebral and cerebellar cortex), cerebral white matter volume, cerebral spinal fluid volume, or hippocampal volume.

A larger, overlapping study of these 8 year old children with the same 3D quantitative volumetric MRI classified MRI findings as normal, mildly abnormal, or severely abnormal and calculated corpus callosum area on a midsagittal T2 slice (Rademaker et al., 2007). There were no differences in presence of brain lesions on

MRI between PT children who were treated with hydrocortisone and those who were not after adjusting for a propensity score representing the likelihood of requiring hydrocortisone. Mean midsagittal corpus callosum area was smaller in the hydrocortisone treated group compared with the non-treated group, but the difference was not significant after adjustment for the propensity score. No other brain differences were identified. This investigation represented the largest group from their sample.

### 3.3 HYDROCORTISONE AND DEXAMETHASONE

Tam and colleagues published a study focused on steroid effects on the cerebrum and cerebellum that examined both hydrocortisone and dexamethasone (Tam et al., 2011). They obtained T2-weighted and T1 3-D volumetric scans at two sites at two time points – as soon after birth as the infant was able and again at term post menstrual age. Adjusting for multiple covariates, there was a decrease in cerebellar growth associated with glucocorticoid exposure resulting in 1.88 cm^3^ smaller cerebellum at 40 weeks post menstrual age after hydrocortisone and 2.31 cm^3^ smaller cerebellum associated with dexamethasone (8% and 10% smaller cerebellar volumes, respectively). They were unable to determine whether there was a dose dependent effect of glucocorticoids on cerebellar growth. Cerebellar volume was not associated with the number of days since the last steroid administration, indicating that the volume changes were not immediately reversible effects. Clinical factors were also associated with decreased cerebellar volume including intubation duration, hypotension, patent ductus arteriosus, but postnatal dexamethasone, postnatal hydrocortisone, and severe intraventricular hemorrhage were associated with the largest decreases in cerebellar volume. The same analyses were performed for cerebral volumes at term. Cerebral volume at term was not associated with postnatal hydrocortisone or dexamethasone exposure.

### 3.4 INHALED GLUCOCORTICOIDS

We identified no studies examining association between inhaled corticosteroids in preterm infants and structural brain outcomes.

## 4.0 DISCUSSION

In this scoping review we identified 11 papers related to associations or effects between postnatal GC and structural or anatomical brain metrics in humans born PT. Dexamethasone was consistently associated with unfavorable brain differences. Radiologist scoring rubrics and quantitative total and regional brain areas or volumes of varied structures were reported. The results showed that exposure to dexamethasone was associated with reduction in volumes across multiple brain regions, including the cerebellum. However, for hydrocortisone, most studies reported no identified differences. Only one study of hydrocortisone exposure reported that the intervention was associated with volumetric differences and these localized to the cerebellum. Overall cohort sizes varied from small to large. The range of treatment dose, duration, and age at initiation varied widely.

### 4.1 GLUCOCORTICOID EFFECTS ON BRAIN DEVELOPMENT

Several lines of evidence support consideration of the effects of GC, and dexamethasone specifically, on brain development in PT infants. GC enter the brain via simple diffusion across the blood brain barrier. Within the brain, GC affect brain development via intracellular glucocorticoid and mineralocorticoid receptors. Glucocorticoid receptor binding causes both inhibition and enhancement of gene transcription (De Kloet et al., 1998; Nishi and Kawata, 2007). Glucocorticoid receptor activation may suppress synaptic plasticity and inhibit neuronal development. Glucocorticoid receptors are widely expressed in the glia and neurons of the cerebrum and cerebellum (Bohn et al., 1994). Mineralocorticoid receptor activation meanwhile may aid in synapse plasticity and neuronal survival (Joëls, 2007; Johnston et al., 2009; McEwen, 1994; McEwen et al., 1992). Mineralocorticoid receptors are primarily found in the hippocampus and limbic structures (Joëls, 2001). Dexamethasone preferentially binds glucocorticoid receptors while hydrocortisone preferentially binds mineralocorticoid receptors. Existing studies have recognized this distribution of glucocorticoid and mineralocorticoid receptors by assessing regional brain volumes, including the cerebellum, and specifically examining hippocampal volumes after hydrocortisone exposure.

Studies in neonatal rats have shown decreased neurogenesis and impaired long term potentiation with dexamethasone, but not hydrocortisone. (Chang, 2014; Cotterrell et al., 1972; Howard and Granoff, 1968; Huang et al., 1999). In the human infant studies we review here, those assessing dexamethasone provide support for the deleterious effects of dexamethasone on brain structure in humans. Early in life, smaller reductions in brain volume were found in studies that used lower cumulative doses and were given later in infancy (Parikh NA et al., 2007) compared with higher doses given earlier (Murphy BP et al., 2001), suggesting dose-dependent and time-sensitive effects. Meanwhile, white matter volume reductions were reported in late adolescence (Cheong et al., 2014), but not at younger ages. This finding at a later age may reflect a selective vulnerability of white matter and altered developmental trajectory that can only be observed over time.

### 4.2 GLUCOCORTICOID EFFECTS ON THE CEREBELLUM

The cerebellum appears to be especially sensitive to GC exposure. Postnatal GC impair rat cerebellar development by decreasing cellular proliferation (Noguchi, 2014).

Similarly, studies of human infants demonstrated decreased cerebellar volume after dexamethasone administration (Parikh NA et al., 2007; Tam et al., 2011). The only study reporting adverse changes after hydrocortisone administration found those differences in the cerebellum (Tam et al., 2011). However, inhibition of cerebellar development may be temporary. The studies of cerebellar volume in childhood and late adolescence did not find differences in cerebellar volume after postnatal dexamethasone administration (Cheong et al., 2014; Lodygensky et al., 2005; Rademaker et al., 2007). It is hypothesized that individuals may have catch up cerebellar growth.

### 4.3 GLUCOCORTICOID EFFECTS ON THE HIPPOCAMPUS

Early GC treatment in rats (postnatal days 2-8) has shown decreased hippocampal glucocorticoid receptor number at pubertal age (postnatal day 40), suggesting lasting impacts on the regulation of cellular development (Zoli et al., 1990). The hippocampus is also important to consider because of the high concentration of mineralocorticoid receptors. (Lodygensky et al., 2005) reports that it was the first study directly addressing the effect of postnatal corticosteroid treatment on hippocampal volumes in humans. The authors assessed hydrocortisone exposure and outcomes at 8 years old. They hypothesized that the absence of differences in their study could be explained by a combination of the decreased potency and shorter half-life of hydrocortisone compared with dexamethasone, and the protective effect of hydrocortisone’s preferential binding of mineralocorticoid receptors. They also pointed to the use of sodium bisulphite as a preservative in dexamethasone as a possible cause of decreased neuronal cell line viability based on in vitro and rodent studies of dexamethasone. Similar to studies of structural brain outcomes at earlier ages, the timing of GC administration later in human neonatal life may have been beyond a particular window of developmental vulnerability. (Cheong et al., 2014) did not identify hippocampal differences in adolescence after dexamethasone exposure. There may be catch up growth in the hippocampus. It may also be that changes in the hippocampus such as changes to glucocorticoid receptor number, are not reflected in volume differences. Future studies should examine metrics other than volume, for instance diffusion metrics, with attention to the hippocampus.

### 4.4 FUTURE DIRECTIONS

The studies we review here highlight the importance of the timing of GC administration and of assessing brain at different ages. The timing of GC administration during the neonatal period could produce different effects on brain structure and development. Infant MRI metrics may serve as proximal biomarkers associated with later developmental outcomes. Future studies should assess advanced MRI metrics as mediators of GC effects on later neurodevelopmental outcomes. Evaluation at later ages is also important to understanding neurodevelopment. GCs in the neonatal period act on a changing substrate. Effects may not be evident until the brain structure is more mature.

Examining subregions or certain types of brain matter (e.g. white vs gray matter; cerebral vs cerebellar), may better identify effects than a whole brain approach. Although not routinely implemented in the clinical setting, no study has yet examined effects of GC using advanced microstructural metrics, for instance diffusion metrics (Feldman et al., 2010) or quantitative microstructural metrics (Mancini et al., 2020). These metrics may get closer to the underlying neurobiology and therefore may detect GC effects that cannot be seen on the more macroscopic volumetric scale that has been reported thus far. Using these quantitative metrics may shed light on contradictory results from the macroscopic volumetric measures.

## 5.0 CONCLUSIONS

GC effects on brain are of interest to a wide audience of researchers across the lifespan and across many clinical conditions. Despite the availability of clinical and research advanced imaging modalities, relatively few human studies have directly assessed the effect of this intervention on brain structural development. This review highlights the need for additional research on neonatal GC and their potential effects on brain development. As GC use becomes more targeted and ongoing research aims to identify optimal populations and treatment regimens, incorporating information about both brain structure in the infant period and in later childhood can provide researchers and clinicians with a better understanding of the downstream effects of this important treatment on neurodevelopment.

## Supporting information

prisma checklist

## Data Availability

All data produced in the present study are available upon reasonable request to the authors

## ACKNOWLEDGEMENTS

This work was supported by the National Institutes of Health (2RO1-HD069150 to Dr. Feldman) and the Young Investigator Award to Dr. Dubner, from the Society of Developmental and Behavioral Pediatrics (2019).

