## Supplemental Material for "Effects of Postnatal Glucocorticoids on Brain Structure in Preterm Infants, A Scoping Review"

**S1. Full Search Term for PubMed**

("infant, newborn"[MeSH Terms] OR "infan*"[Text Word] OR "neonat*"[Text Word] OR "newborn*"[Text Word] OR "premature"[Text Word] OR "low birth weight"[Text Word] OR "VLBW"[Text Word] OR "LBW"[Text Word] OR "ELBW"[Text Word] OR ("Premature birth"[MeSH Terms] OR "infant, premature"[MeSH Terms] OR "infant, very low birth weight"[MeSH Terms] OR "infant, premature, diseases"[MeSH Terms] OR (("preterm"[Text Word] OR "premature"[Text Word] OR "Prematurity"[Text Word] OR "prematurely"[Text Word]) AND ("deliver*"[Text Word] OR "birth*"[Text Word] OR "infant*"[Text Word] OR "infancy"[Text Word] OR "baby"[Text Word] OR "babies"[Text Word] OR "neonat*"[Text Word] OR "newborn*"[Text Word] OR "perinatal"[Text Word] OR "parturition"[Text Word])) OR "preemi*"[Text Word] OR "premi"[Text Word] OR "premie"[Text Word] OR "premies"[Text Word] OR "preterm"[Text Word] OR "low birth weight"[Text Word] OR "low birthweight"[Text Word] OR "early gestation*"[Text Word] OR "low gestation*"[Text Word])) AND ("Brain"[MeSH Terms] OR "Brain"[Text Word] OR "nerve fibers, myelinated"[MeSH Terms] OR "white matter*"[Text Word] OR "myelinated nerve fiber*"[Text Word] OR "Glia"[Text Word] OR "gray matter"[Text Word] OR "cerebel*"[Text Word] OR "ventric*"[Text Word] OR "cerebrum"[Text Word] OR "corpus callosum"[Text Word] OR "internal capsules"[Text Word] OR "external capsules"[Text Word] OR "anterior commissure"[Text Word] OR "limbic system"[Text Word]) AND ("hydrocortisone"[MeSH Terms] OR "dexamethasone"[MeSH Terms] OR "betamethasone"[MeSH Terms] OR "steroids"[MeSH Terms] OR "Glucocorticoids"[MeSH Terms] OR "Adrenal Cortex Hormones"[MeSH Terms] OR "hydrocortisone*"[Text Word] OR "betamethason*"[Text Word] OR "dexamethason*"[Text Word] OR "steroid*"[Text Word] OR "corticoster*"[Text Word] OR "corticoid*"[Text Word] OR "glucocortic*"[Text Word] OR "adrenal cortex hormone*"[Text Word]) AND "English"[Language]

**S2. Studies of glucocorticoids that reported a brain injury as a clinical characteristic.**

We identified 50 studies that reported a brain injury outcome, most commonly IVH, as a characteristic of the study population after treatment with corticosteroid, but did not directly assess the effect of the steroid on brain injury. These studies are listed in the **Table**. (Sinkin et al., 2000) is a representative example of this category. Sinkin and colleagues conducted a multi-center, double blind phase 2 randomized controlled trial of a two dose course of dexamethasone early in neonatal life to assess the clinical efficacy and safety of the medication. The study population included infants under 30 weeks gestation with respiratory distress syndrome who required mechanical ventilation at 12-18 hours of life after surfactant. The dosing regimen was 0.5 mg dexamethasone administered via IV at 12-18 hours of life and 12 hours later. Later dexamethasone use was permitted per clinician discretion. The relevant brain outcome was presence and severity of IVH. 384 infants were enrolled and 189 received dexamethasone. There was no difference between the groups in incidence or severity of intraventricular hemorrhage. Of note, LeFlore (LeFlore JL et al., 2002) reported on an investigation of infants who received either no treatment, antenatal dexamethasone, postnatal dexamethasone only, or both antenatal and postnatal steroids. However, the aim of the study and the reported results were on the effect of the antenatal exposure on cranial ultrasound at discharge. They report that the likelihood of abnormal ultrasound at discharge was greater in the postnatal dexamethasone groups compared with the no-postnatal dexamethasone groups. They do not report on the relationship between postnatal dexamethasone and specific cranial ultrasound findings as they do for antenatal dexamethasone at discharge.

**S3. Glucocorticoids as risk factors**

12 studies examined perinatal clinical risk factors and their effect on brain development (Batton et al., 2012; Cuzzilla R et al., 2018; Heo et al., 2018; Kelly CE et al., 2014; Kidokoro H et al., 2014; Matthews LG et al., 2018; Neubauer et al., 2017; Parikh NA et al., 2021; Thompson DK et al., 2008; Thompson et al., 2012; Vesoulis ZA et al., 2018; Watterberg et al., 2004). For the most part, steroid type and administration details are not specified. Whether or not steroid administration functions as a proxy for some other factor, such as pulmonary health, was not able to be discerned.

**Table**. Studies reporting brain injury as an outcome

| **Steroid** | **Imaging** | **Author Year** | **Study Type** | **Number of Participants** | **Administration Details** | **Brain outcome** | **Result** |
| --- | --- | --- | --- | --- | --- | --- | --- |
| Dexamethasone | US | (Doyle et al., 2006) | RCT | 70 | 0.15 mg/kg/d for 3d  0.10 mg/kg/d for 3d  0.05 mg/kg/d for 2d  0.02 mg/kg/d for 2d  Total of 0.89 mg/kg over 10 days, intravenously | -Intraparenchymal Hemorrhage  -Cystic PVL | No association |
|  | US | (Gaissmaier and Pohlandt, 1999) | RCT | 17 | 0.25mg/kg, intravenously  Children who at completion of the study period were still receiving epinephrine received DEX (0.25 mg/kg) as a rescue dose.  Age at first dose: 2d (1-20) | -IVH (Grade III-IV) | No association |
|  | US | (LeFlore JL et al., 2002) | Cohort Study | 173 | 0.3 – 0.5 mg/kg tapered over 14- to 42 days  Age at first dose:  30 ± 14 days (no antenatal glucocorticoids)  25 ± 10 days (plus antenatal glucocorticoids) | Abnormal cranial ultrasound at discharge | Higher likelihood in infants exposed to postnatal dexamethasone |
|  | US | (Lim G et al., 2015) | Cohort Study | 131 | 3.18 mg/kg, 14d course  1.10 mg/kg, 10d course  1.59 mg/kg, 7d course  Age at first dose: 21d or later | Severe-grade IVH/cystic PVL | No association |
|  | US | (Lin et al., 1999) | RCT | 40 | 1 dose q12hr, intravenously  0.25 mg/kg/dose, days 1-7  0.12 mg/kg/dose, days 8-14  0.05 mg/kg/dose, days 15-21  0.02 mg/kg/dose, days 22-28  Mean age at first dose: 8.0 ± 4.6 hrs. | -IVH (>Grade II) | No association |
|  | US | (Malloy et al., 2005) | RCT | 16 | Low dose: 0.08 mg/kg/d for 7d  High dose: 0.5 mg/kg/d for 3d followed by 0.3mg/kg/d for 4d.  The daily dose for each group was divided into 2 doses q12hr, intravenously | -Mild, moderate, and severe ventricular hypertrophy | No association |
|  | US | (McEvoy et al., 2004) | RCT | 62 | High dose: 0.5 mg/kg/d for 3d, 0.25 mg/kg/d for 3d, 0.1 mg/kg/d on day 7. Total dose: 2.35 mg/kg  Low dose: 0.2 mg/kg/d for 3d, 0.1 mg/kg/d for 4d. Total dose: 1mg/kg,  All daily doses given q12hr, intravenously. | -PVL | No association |
|  | US | (C. Romagnoli et al., 2002) | RCT | 50 | 0.5 mg/kg/d for 3d  0.25 mg/kg/d for 3d  0.125 mg/kg/d on the 7th day  Intravenously  Age at first dose: 4d | -Major cranial US abnormalities at 12 months of corrected age.  -Periventricular Leukomalacia. | No association |
|  | US | (Sanders et al., 1994) | RCT | 40 | 0.5 mg/kg, 2 doses q12hrs, intravenously  Age at first dose: 12-18hrs. | -IVH (Grade I) | No association |
|  | US | (Subhedar et al., 1997) | RCT | 84 | 0.5 mg/kg/dose, 6 doses.  0.25 mg/kg/dose, 6 doses.  6d total, intravenously. | -IVH | Infants with an existing IVH were included in this study. Progression was detected only in control infants. |
|  | US | (Sinkin et al., 2000) | RCT | 384 | 0.5 mg/kg with the first dose administered between 12and 18hrs of age and the second dose 12hrs later, intravenously. | -IVH | No association |
|  | US | (Stark et al., 2001) | RCT | 220 | 0.15 mg/kg/d for 3d  0.10 mg/kg for 3d  0.05 mg/kg for 2d  0.02 mg/kg for 2d  Intravenously or orally  Age at first dose: 14.2 ± 5.8hrs | -IVH (Any grade)  -Periventricular Leukomalacia | No association |
|  | US | (Suske et al., 1996) | RCT | 26 | 0.5 mg/kg, divided into 2 doses for 5d, intravenously  Surfactant: 100mg/kg  Age at first dose: <2hrs | -ICH (Grade I-II) | No association |
|  | US | (Tsukahara et al., 1999) | RCT | 26 | 0.125 mg/kg, 6 doses q12hrs, intravenously  Age at first dose: evening of day 4. | -Intracranial hemorrhage  -Periventricular Leukomalacia | No association |
|  | US | (Yaseen et al., 1999) | RCT | 29 | 0.5mg/d for 2d  0.3 mg/kg/d for 2d  0.2 mg/kg/d on the 5th day of treatment,  Intravenously  Age at first dose 2.8 ± 1.01 hrs. | -IVH | No association |
|  | US | (Yeh et al., 1990) | RCT | 57 | 1 dose q12hrs, intravenously  0.5 mg/kg/dose, days 1-3  0.25 mg/kg/dose, days 4-6  0.12 mg/kg/dose, days 7-9  0.05 mg/kg/dose, days 10-12  Age at first dose: 8.5 ± 3.1 hrs. | -IVH | No association |
|  | US | (Yeh et al., 1997) | RCT | 262 | 1 dose q12hrs, intravenously  0.25 mg/kg/dose, days 1-7  0.12 mg/kg/dose, days 8-14  0.05 mg/kg/dose, days 15-21  0.02 mg/kg/dose, days 22-28  Age at first dose: 7.4 ± 5.2 hrs. | -IVH (Grade <II) | No association |
|  | US | (Yeh et al., 1998) | RCT | 133 | 0.25 mg/kg/dose bid, days 1-7  0.12 mg/kg/dose bid, days 8-14  0.05 mg/kg/dose bid, days 15-21  0.02 mg/kg/dose bid, days 22-28  Intravenously. Age at first dose: within 12hrs after birth | -IVH (Grade >II) | No association |
|  | US | (Anttila E et al., 2005) | RCT | 109 | 0.25 mg/kg, 4 doses, 2d q12hrs.  Age at first dose: 6hrs. | -ICH (Grade III-IV)  -PVL | No association |
|  | US | (Brozanski et al., 1995) | RCT | 78 | 0.25 mg/kg/dose twice daily for 3d,  Intravenously, if IV not possible IM  Age at first dose: 7d postnatal age. | -IVH (Grade <II) | Decreased incidence of worsening IVH |
|  | US | (Durand et al., 1995) | RCT | 43 | 0.5mg/kg/d, 2 divided doses for the first 3d  0.25 mg/kg/d for the next 3d  0.1 mg/kg/d on the 7th day of treatment,  Intravenously  Age at first dose: 7-14d | -IVH | No association |
|  | US | (Garland et al., 1999) | RCT | 241 | Total dose:  1.35 mg/kg over 3d (n=41)  1.175 mg/kg over 3d (n=77)  3-day tapering course q12hrs.  Age at first dose: starting at 24 to 48hrs of life. | -ICH (Any grade)  -PVL | No association |
|  | US | (Kopelman AE et al., 1999) | RCT | 70 | 0.2mg/kg, 1 dose, intravenously  Age at first dose: within 2hrs of delivery. | -IVH (Grade III-IV) | Study was terminated early because they were underpowered to detect differences in IVH.  DEX group showed trends toward grade III-IV IVH and death. |
|  | US | (Mieskonen et al., 2003) | RCT | 16 | 0.5 mg/kg/d, divided into 2 doses  Age at first dose: 28d. | -IVH | No association. |
|  | US | (Odd et al., 2004) | RCT | 33 | Individual course:  0.5mg/kg/d for 3d, 0.3mg/kg/d for 3d, 0.1mg/kg/d for 3d, then 0.1mg/kg every 72hr until the infant was extubated and required a FiO2 < 0.25 for three doses 9d). If there was clinical deterioration, the dose reverted to 0.3mg/kg/d for 3 days, then was weaned again using the same protocol.  Total dose of 3.8 (2.0-5.7) mg/kg.  42d course:  0.5mg/kg/d for 3d, 0.3mg/kg/d for 3d, then a dose decreasing by 10% every 3d to 0.1mg/kg/d over a further 30d, then 0.1mg/kg on alternate days for one further week.  Total dose of 6.5 (3.8-7.3) mg/kg. | -IVH (Grade III-IV) | No association |
|  | US | (Rastogi et al., 1996) | RCT | 70 | 0.5mg/kg/d for 12d, intravenously  Age at first dose: 7.2 ± 3hrs postnatal age. | -IVH (Any grade) | No association |
|  | US | (Romagnoli C et al., 1999) | RCT | 50 | 0.5 mg/kg/d for 3d  0.25 mg/kg/d for 3d  0.125 mg/kg/d on the 7th day of treatment,  Intravenously  Age at first dose: 4d. | -ICH (Grade >II) | No association |
|  | US | (C Romagnoli et al., 2002) | RCT | 30 | 0.5 mg/kg/d for 6d  0.25 mg/kg/d for 6d  0.125 mg/kg/d for 2d  Total cumulative dose of 4.75 mg/kg, intravenously  Age at first dose: 10d. | -Major cranial US abnormalities at 12 months of corrected age.  -Cystic PVL  -IVH (Grade >II) | No association |
|  | US | (Shinwell et al., 1996) | RCT | 248 | 0.25 mg/kg, 6 doses q12hrs, intravenously  Age at first dose: 1d. | -IVH (Garde <II)  -PVL | No association |
|  | US | (Shinwell et al., 1999) | RCT | 159 | 0.25 mg/kg, 6 doses q12hrs, intravenously  Age at first dose: <12hrs. | -IVH (Grade I-IV)  -PVL | No association. |
|  | US | (Smets and Schwagten, 2000) | Case Control Study | 48 | 0.125 mg/kg for 3d. | -Germinolysis | No association |
|  | US | (Soll, 1999) | RCT | 542 | 0.5 mg/kg/d for 3d  0.25mg/kg/d for 3d  0.10 mg/kg/d for 3d  0.05 mg/kg/d for 3d  Intravenously  Age at first dose: 12hrs. | -IVH (Any grade)  -PVL | No association |
|  | US | (Tapia et al., 1998) | RCT | 109 | 0.5mg/kg/d, days 1-3  0.25 mg/kg/d, days 4-6  0.12 mg/kg/d, days 7-9  0.06 mg/kg/d, days 10-12  Intravenously | -IVH | No association |
|  | US | (Yates et al., 2019) | RCT | 22 | 50 µg/kg, once daily on days 1-10 after randomization (i.e.,10 doses), then given on alternate days (i.e., on days 12, 14 and 16), making a total of 13 doses.  Intravenously, nasogastric tube or orally.  Total cumulative dose of 0.65 mg/kg. | -IVH (Any grade)  -Hydrocephalus  -PVL  -Other white-matter injury | Determined it to not be feasible to conduct this trial due to low levels or recruitment. |
| Hydrocortisone | US | (Biswas et al., 2003) | RCT | 253 | 1 mg/kg/d for days 1-4, then halved on day 5. Same dose days 5-7, intravenously  Age at first dose: 5hrs. | -IVH (Grade III-IV)  -PVL | No association |
|  | US | (Bonsante et al., 2007) | RCT | 50 | 0.5 mg/kg, q12hr for 9d  0.5 mg/kg/d for 3d  Intravenously  Age at first dose: <48hrs of life. | -IVH (Grade III)  -PVL | No association |
|  | US | (Bourchier and Weston, 1997) | RCT | 40 | 2.5 mg/kg, 2 doses, 4hrs apart. Subsequent doses were q6hr for the remainder of the treatment period. The initial dose (2.5 mg/kg) was continued for 48hrs, followed by 1.25 mg/kg  for 48hrs, then 0.625 mg/kg for a further 48hrs before stopping treatment, intravenously  Age at first dose: 11.4hrs. | -IVH (grades II-IV) | No association |
| Hydrocortisone | US | (Efird et al., 2005) | RCT | 34 | 1mg/kg BID for 2d  0.3 mg/kg BID for 3d  Intravenously | -IVH (Grade >II)  -PVL | No association |
|  | US | (Ng et al., 2006) | RCT | 48 | 1 mg/kg/dose, q8hr for 5d, intravenously | -IVH (Grade >III)  -PVL | No association |
|  | MRI/US | (Parikh et al., 2015) | RCT | 57 | q12h, 3mg/kg/d for 4 days, 2mg/kg/d for 2d, 1mg/kg/d for 1d.  Total cumulative dose of 17 mg/kg, intravenously | -White matter injury near 36 weeks age (cystic  periventricular leukomalacia, porencephalic cyst, parenchymal hemorrhage, and/or ventriculomegaly (with or without intraventricular hemorrhage) | No association |
|  | US | (Verma RP et al., 2017) | Cohort Study | 143 | 2-4 mg/kg/d, intravenously | -Ventriculomegaly (VM)  -IVH | HC group had higher risk for IVH which declined in the multivariate analysis.  A trend towards lower risk of VM was noted in HC group, which became significant after controlling for body weight. |
|  | US | (Watterberg et al., 2007) | RCT | 252 | 1 mg/kg/d (8 –10mg/m2/d) divided twice a day for 12d. Then 0.5 mg/kg/d for 3d | -IVH (Grade II-IV)  -PVL | No association |
|  | MRI | (Takayanagi et al., 2015) | Cohort Study | 58 | Intravenously: 9.0 ± 7.2 (1.0-28.2) mg/kg  Orally: 68.1 ± 34.1 (0-161.5) mg/kg.  Mean duration of 57.1 ± 27.8 (0-154) d. | MRI abnormality (focal high intensity of white matter or irregular ventricular wall with mild dilation) | No difference in hydrocortisone dose between patients with and without MRI abnormality |
|  | MRI, US | (Baud et al., 2016) | RCT | 521 | 1mg/kg/d, 2 doses for 7d  0.5mg/kg/d, 1 dose for 3d  Intravenously | -IVH (Grade III-IV)  -Cystic PVL | No association |
|  | US | (Onland et al., 2019) | RCT | 372 | 5 mg/kg/d, 4 doses/d for 7d  3.75 mg/kg/d, 3 doses/d for 5d, subsequently lowering the frequency by 1 dose every 5 days.  Cumulative dose of 72.5 mg/kg.  Age at first dose: between 7-14d 3.75 mg/kg/d, 3 doses/d for 5d, subsequently lowering the frequency by 1 dose every 5 days.  Cumulative dose: 72.5 mg/kg. | -IVH (Grade >II)  -PVL | No association |
|  | US | (Peltoniemi et al., 2005) | RCT | 51 | 2.0 mg/kg, divided into 3 doses q8hrs for 2d  1.5 mg/kg, divided into 3 doses q8hrs for 2d  0.75 mg/kg, divided into 2 doses q12hrs for 6d  Intravenously  Age at first dose: before the age of 36hrs. | -IVH (Any grade)  -Cystic PVL | No association |
| Betamethasone | US | (Smolkin et al., 2014) | Cohort Study | 35 | 0.1 mg/kg/dose, twice daily for 3d  0.05 mg/kg/dose twice daily for 2d  0.05 mg/kg/dose once daily for 2d  Orally (PO)  Mean age at first dose: 31.0 ± 13.6 d. | -PVL | No association |
| Budesonide | US | (Bassler et al., 2015) | RCT | 856 | Two inhaled puffs of 200 μg each q12hr in the first 14d of life and one puff q12hr from day 15 until the last dose of the study drug had been administered.  Age at first dose: 12hrs after random assignment. | -Brain injury | No association |
| Dexamethasone and Budesonide | US | (Kovács et al., 1998) | RCT | 60 | DEX (IV): 0.25 mg/kg, q12hr for 3d  BUD (INH): 500mg, twice daily for 18d  Age at first dose: 7d. | -IVH (Any grade) | No association |
|  | US | (Halliday et al., 2001) | RCT | 570 | DEX (IV):  0.50mg/kg/d, 2 divided doses for 3d  0.25mg/kg/d, 2 divided doses for 3d  0.10mg/kg/d, 2 divided doses for 3d  0.05mg/kg/d, 2 divided doses for 3d  BUD (INH): A 500- to 1000-g infant was given 400mg (2 puffs) twice daily and a 1000- to 1500-g infant 600mg (3 puffs) twice daily.  Age at first dose:  Early cohort: <72 hours  Delayed cohort: >15 days | - major cerebral abnormality  -IVH | No association |

Anttila E, Peltoniemi O, Haumont D, Herting E, ter Horst H, Heinonen K, Kero P, Nykänen P, Oetomo SB, Hallman M, 2005. Early neonatal dexamethasone treatment for prevention of bronchopulmonary dysplasia. Randomised trial and meta-analysis evaluating the duration of dexamethasone therapy. Eur J Pediatr 164, 472–81. https://doi.org/10.1007/s00431-005-1645-8

Bassler, D., Plavka, R., Shinwell, E.S., Hallman, M., Jarreau, P.-H., Carnielli, V., Van den Anker, J.N., Meisner, C., Engel, C., Schwab, M., Halliday, H.L., Poets, C.F., 2015. Early Inhaled Budesonide for the Prevention of Bronchopulmonary Dysplasia. N Engl J Med 373, 1497–1506. https://doi.org/10.1056/NEJMoa1501917

Batton, B.J., Li, L., Newman, N.S., Das, A., Watterberg, K.L., Yoder, B.A., 2012. Eunice Kennedy Shriver National Institute of Child Health and Human Development Neonatal Research Network. Feasibility study of early blood pressure management in extremely preterm infants. Journal of Pediatrics 161, 65–9. https://doi.org/10.1016/j.jpeds.2012.01.014

Baud, O., Maury, L., Lebail, F., Ramful, D., El Moussawi, F., Nicaise, C., Zupan-Simunek, V., Coursol, A., Beuchée, A., Bolot, P., Andrini, P., Mohamed, D., Alberti, C., PREMILOC trial study group, 2016. Effect of early low-dose hydrocortisone on survival without bronchopulmonary dysplasia in extremely preterm infants (PREMILOC): a double-blind, placebo-controlled, multicentre, randomised trial. Lancet 387, 1827–1836. https://doi.org/10.1016/S0140-6736(16)00202-6

Biswas, S., Buffery, J., Enoch, H., Bland, M., Markiewicz, M., Walters, D., 2003. Pulmonary effects of triiodothyronine (T3) and hydrocortisone (HC) supplementation in preterm infants less than 30 weeks’ gestation: results of the THORN trial - thyroid hormone replacement in neonates. Pediatric Research 53, 48–56. https://doi.org/10.1203/00006450-200301000-00011

Bonsante, F., Latorre, G., Iacobelli, S., Forziati, V., Laforgia, N., Esposito, L., 2007. Early low-dose hydrocortisone in very preterm infants: a randomized, placebo-controlled trial. Neonatology 91, 217–21. https://doi.org/10.1159/000098168

Bourchier, D., Weston, P.J., 1997. Randomised trial of dopamine compared with hydrocortisone for the treatment of hypotensive very low birthweight infants. Arch Dis Child Fetal Neonatal Ed 76, F174-178. https://doi.org/10.1136/fn.76.3.f174

Brozanski, B., Jones, J., Gilmour, C., Balsan, M., Vazquez, R., Israel, B., Newman, B., Mimouni, F., Guthrie, R., 1995. Effect of pulse dexamethasone therapy on the incidence and severity of chronic lung disease in the very low birth weight infant. Journal of pediatrics 126, 769‐776. https://doi.org/10.1016/s0022-3476(95)70410-8

Cuzzilla R, Spittle AJ, Lee KJ, Rogerson S, Cowan FM, Doyle LW, Cheong JLY, 2018. Postnatal Brain Growth Assessed by Sequential Cranial Ultrasonography in Infants Born <30 Weeks’ Gestational Age. AJNR Am J Neuroradiol 39, 1170–1176. https://doi.org/10.3174/ajnr.A5679

Doyle, L.W., Davis, P.G., Morley, C.J., McPhee, A., Carlin, J.B., 2006. Low-dose dexamethasone facilitates extubation among chronically ventilator-dependent infants: a multicenter, international, randomized, controlled trial. Pediatrics 117, 75–83. https://doi.org/10.1542/peds.2004-2843

Durand, M., Sardesai, S., McEvoy, C., 1995. Effects of early dexamethasone therapy on pulmonary mechanics and chronic lung disease in very low birth weight infants: a randomized, controlled trial. Pediatrics 95, 584‐590.

Efird, M.M., Heerens, A.T., Gordon, P.V., Bose, C.L., Young, D.A., 2005. A randomized-controlled trial of prophylactic hydrocortisone supplementation for the prevention of hypotension in extremely low birth weight infants. Journal of Perinatology 25, 119–24. https://doi.org/10.1038/sj.jp.7211193

Gaissmaier, R.E., Pohlandt, F., 1999. Single-dose dexamethasone treatment of hypotension in preterm infants. Journal of Pediatrics 134, 701–5. https://doi.org/10.1016/s0022-3476(99)70284-2

Garland, J.S., Alex, C.P., Pauly, T.H., Whitehead, V.L., Brand, J., Winston, J.F., 1999. A three-day course of dexamethasone therapy to prevent chronic lung disease in ventilated neonates: a randomized trial. Pediatrics 104, 91–9. https://doi.org/10.1542/peds.104.1.91

Halliday, H.L., Patterson, C.C., Halahakoon, C.W., 2001. A multicenter, randomized open study of early corticosteroid treatment (OSECT) in preterm infants with respiratory illness: comparison of early and late treatment and of dexamethasone and inhaled budesonide. Pediatrics 107, 232–40.

Heo, J.S., Kim, E.-K., Choi, Y.H., Shin, S.H., Sohn, J.A., Cheon, J.-E., Kim, H.-S., 2018. Timing of sepsis is an important risk factor for white matter abnormality in extremely premature infants with sepsis. Pediatr Neonatol 59, 77–84. https://doi.org/10.1016/j.pedneo.2017.07.008

Kelly CE, Cheong JL, Molloy C, Anderson PJ, Lee KJ, Burnett AC, Connelly A, Doyle LW, Thompson DK, 2014. Neural correlates of impaired vision in adolescents born extremely preterm and/or extremely low birthweight. PLoS One 9, e93188. https://doi.org/10.1371/journal.pone.0093188

Kidokoro H, Anderson PJ, Doyle LW, Woodward LJ, Neil JJ, Inder TE, 2014. Brain injury and altered brain growth in preterm infants: predictors and prognosis. Pediatrics 134, e444-53. https://doi.org/10.1542/peds.2013-2336

Kopelman AE, Moise AA, Holbert D, Hegemier SE, 1999. A single very early dexamethasone dose improves respiratory and cardiovascular adaptation in preterm infants. J Pediatr 135, 345–50. https://doi.org/10.1016/s0022-3476(99)70132-0

Kovács, L., Davis, G., Faucher, D., Papageorgiou, A., 1998. Efficacy of sequential early systemic and inhaled corticosteroid therapy in the prevention of chronic lung disease of prematurity. Acta paediatrica 87, 792‐798. https://doi.org/10.1080/080352598750013905

LeFlore JL, Salhab WA, Broyles RS, Engle WD, 2002. Association of antenatal and postnatal dexamethasone exposure with outcomes in extremely low birth weight neonates. Pediatrics 110, 275–9. https://doi.org/10.1542/peds.110.2.275

Lim G, Lee BS, Choi YS, Park HW, Chung ML, Choi HJ, Kim EA, Kim KS, 2015. Delayed Dexamethasone Therapy and Neurodevelopmental Outcomes in Preterm Infants with Bronchopulmonary Dysplasia. Pediatr Neonatol 56, 261–7. https://doi.org/10.1016/j.pedneo.2014.11.006

Lin, Y.J., Yeh, T.F., Hsieh, W.S., Chi, Y.C., Lin, H.C., Lin, C.H., 1999. Prevention of chronic lung disease in preterm infants by early postnatal dexamethasone therapy. Pediatric Pulmonology 27, 21–6. https://doi.org/10.1002/(sici)1099-0496(199901)27:1

Malloy, C.A., Hilal, K., Weiss, M.G., Rizvi, Z., Muraskas, J.K., 2005. A prospective, randomized, double-masked trial comparing low dose to conventional dose dexamethasone in neonatal chronic lung disease. Internet Journal of Pediatrics and Neonatology 5.

Matthews LG, Inder TE, Pascoe L, Kapur K, Lee KJ, Monson BB, Doyle LW, Thompson DK, Anderson PJ, 2018. Longitudinal Preterm Cerebellar Volume: Perinatal and Neurodevelopmental Outcome Associations. Cerebellum 17, 610–627. https://doi.org/10.1007/s12311-018-0946-1

McEvoy, C., Bowling, S., Williamson, K., McGaw, P., Durand, M., 2004. Randomized, double-blinded trial of low- dose dexamethasone: II. Functional residual capacity and pulmonary outcome in very low birth weight infants at risk for bronchopulmonary dysplasia. Pediatric Pulmonology 38, 55–63.

Mieskonen, S., Eronen, M., Malmberg, L., Turpeinen, M., Kari, M., Hallman, M., 2003. Controlled trial of dexamethasone in neonatal chronic lung disease: an 8-year follow-up of cardiopulmonary function and growth. Acta paediatrica 92, 896‐904.

Neubauer, V., Djurdjevic, T., Griesmaier, E., Biermayr, M., Gizewski, E.R., Kiechl-Kohlendorfer, U., 2017. Routine Magnetic Resonance Imaging at Term-Equivalent Age Detects Brain Injury in 25% of a Contemporary Cohort of Very Preterm Infants. PLoS ONE 12, e0169442. https://doi.org/10.1371/journal.pone.0169442

Ng, P.C., Lee, C.H., Bnur, F.L., Chan, I.H., Lee, A.W., Wong, E., 2006. A double-blind, randomized, controlled study of a “stress dose” of hydrocortisone for rescue treatment of refractory hypotension in preterm infants. Pediatrics 117, 367–75. https://doi.org/10.1542/peds.2005-0869

Odd, D.E., Armstrong, D.L., Teele, R.L., Kuschel, C.A., Harding, J.E., 2004. A randomized trial of two dexamethasone regimens to reduce side-effects in infants treated for chronic lung disease of prematurity. Journal of Paediatrics and Child Health 40, 282–9. https://doi.org/10.1111/j.1440-1754.2004.00364.x

Onland, W., Cools, F., Kroon, A., Rademaker, K., Merkus, M.P., Dijk, P.H., 2019. Effect of hydrocortisone therapy initiated 7 to 14 days after birth on mortality or bronchopulmonary dysplasia among very preterm infants receiving mechanical ventilation: a randomized clinical trial. JAMA 321, 354–63. https://doi.org/10.1001/jama.2018.21443

Parikh, N., Kennedy, K., Lasky, R., Tyson, J., 2015. Neurodevelopmental Outcomes of Extremely Preterm Infants Randomized to Stress Dose Hydrocortisone. PloS one 10, e0137051. https://doi.org/10.1371/journal.pone.0137051

Parikh NA, Sharma P, He L, Li H, Altaye M, Priyanka Illapani VS, 2021. Perinatal Risk and Protective Factors in the Development of Diffuse White Matter Abnormality on Term-Equivalent Age Magnetic Resonance Imaging in Infants Born Very Preterm. J Pediatr 233, 58-65.e3. https://doi.org/10.1016/j.jpeds.2020.11.058

Peltoniemi, O., Kari, M.A., Heinonen, K., Saarela, T., Nikolajev, K., Andersson, S., 2005. Pretreatment cortisol values may predict responses to hydrocortisone administration for the prevention of bronchopulmonary dysplasia in high-risk infants. Journal of Pediatrics 146, 632–7. https://doi.org/10.1016/j.jpeds.2004.12.040

Rastogi, A., Akintorin, S.M., Bez, M.L., Morales, P., Pildes, R.S., 1996. A controlled trial of dexamethasone to prevent bronchopulmonary dysplasia in surfactant-treated infants. Pediatrics 98, 204–10.

Romagnoli, C., Zecca, E., Luciano, R., Torrioli, G., Tortorolo, G., 2002. Controlled trial of early dexamethasone treatment for the prevention of chronic lung disease in preterm infants: a 3-year follow-up. Pediatrics 109. https://doi.org/10.1542/peds.109.6.e85

Romagnoli, C, Zecca, E., Luciano, R., Torrioli, G., Tortorolo, G., 2002. A three year follow-up of preterm infants after moderately early treatment with dexamethasone. Archives of disease of childhood fetal and neonatal edition 87, F55‐8.

Romagnoli C, Zecca E, Vento G, De Carolis MP, Papacci P, Tortorolo G, 1999. Early postnatal dexamethasone for the prevention of chronic lung disease in high-risk preterm infants. Intensive Care Med 25, 717–21. https://doi.org/10.1007/s001340050935

Sanders, R.J., Cox, C., Phelps, D.L., Sinkin, R.A., 1994. Two doses of early intravenous dexamethasone for the prevention of bronchopulmonary dysplasia in babies with respiratory distress syndrome. Pediatric Research 36, 122–8. https://doi.org/10.1203/00006450-199407001-00022

Shinwell, E., Karplus, M., Reich, D., 1999. Early dexamethasone therapy is associated with increased incidence of cerebral palsy 240‐254.

Shinwell, E.S., Karplus, M., Zmora, E., Reich, D., Rothschild, A., Blazer, S., 1996. Failure of early postnatal dexamethasone to prevent chronic lung disease in infants with respiratory distress syndrome. Archives of Disease in Childhood. Fetal and Neonatal Edition 74. https://doi.org/10.1136/fn.74.1.f33

Sinkin, R.A., Dweck, H.S., Horgan, M.J., Gallaher, K.J., Cox, C., Maniscalco, W.M., 2000. Early dexamethasone - attempting to prevent chronic lung disease. Pediatrics 105, 542–8. https://doi.org/10.1542/peds.105.3.542

Smets, K., Schwagten, B., 2000. Postnatal cystic germinolysis and neonatal chronic lung disease: evaluation of risk factors and neurodevelopmental outcome. Acta Paediatr 89, 1111–1114. https://doi.org/10.1080/713794556

Smolkin, T., Ulanovsky, I., Jubran, H., Blazer, S., Makhoul, I.R., 2014. Experience with oral betamethasone in extremely low birthweight infants with bronchopulmonary dysplasia. Archives of Disease in Childhood. Fetal and Neonatal Edition 99. https://doi.org/10.1136/archdischild-2014-306619

Soll, R.F., 1999. Vermont Oxford Network Steroid Study Group. Early postnatal dexamethasone therapy for the prevention of chronic lung disease. Pediatric Research 45.

Stark, A.R., Carlo, W.A., Tyson, J.E., Papile, L.A., Wright, L.L., Shankaran, S., 2001. National Institute of Child Health and Human Development Neonatal Research Network. Adverse effects of early dexamethasone in extremely-low-birth-weight infants. New England Journal of Medicine 344, 95–101. https://doi.org/10.1056/NEJM200101113440203

Subhedar, N.V., Ryan, S.W., Shaw, N.J., 1997. Open randomised controlled trial of inhaled nitric oxide and early dexamethasone in high risk preterm infants. Archives of Disease in Childhood. Fetal and Neonatal Edition 77. https://doi.org/10.1136/fn.77.3.f185

Suske, G., Oestreich, K., Varnholt, V., Lasch, P., Kachel, W., 1996. Influence of early postnatal dexamethasone therapy on ventilator dependency in surfactant-substituted preterm infants. Acta Paediatrica 85, 713–8. https://doi.org/10.1111/j.1651-2227.1996.tb14132.x

Takayanagi, T., Matsuo, K., Egashira, T., Mizukami, T., 2015. Neonatal hydrocortisone therapy does not have a serious suppressive effect on the later function of the hypothalamus-pituitary-adrenal axis. Acta Paediatr. 104, e195-199. https://doi.org/10.1111/apa.12926

Tapia, J.L., Ramirez, R., Cifuentes, J., Fabres, J., Hubner, M.E., Bancalari, A., 1998. The effect of early dexamethasone administration on bronchopulmonary dysplasia in preterm infants with respiratory distress syndrome. Journal of Pediatrics 132, 48–52. https://doi.org/10.1016/s0022-3476(98)70483-4

Thompson, D.K., Inder, T.E., Faggian, N., Warfield, S.K., Anderson, P.J., Doyle, L.W., Egan, G.F., 2012. Corpus callosum alterations in very preterm infants: perinatal correlates and 2 year neurodevelopmental outcomes. Neuroimage 59, 3571–81. https://doi.org/10.1016/j.neuroimage.2011.11.057

Thompson DK, Wood SJ, Doyle LW, Warfield SK, Lodygensky GA, Anderson PJ, Egan GF, Inder TE, 2008. Neonate hippocampal volumes: prematurity, perinatal predictors, and 2-year outcome. Ann Neurol 63, 642–51. https://doi.org/10.1002/ana.21367

Tsukahara, H., Watanabe, Y., Yasutomi, M., Kobata, R., Tamura, S., Kimura, K., 1999. Early (4-7 days of age) dexamethasone therapy for prevention of chronic lung disease in preterm infants. Biology of the Neonate 76, 283–90. https://doi.org/10.1159/000014170

Verma RP, Dasnadi S, Zhao Y, Chen HH, 2017. A comparative analysis of ante- and postnatal clinical characteristics of extremely premature neonates suffering from refractory and non-refractory hypotension: Is early clinical differentiation possible? Early Hum Dev 113, 49–54. https://doi.org/10.1016/j.earlhumdev.2017.07.010

Vesoulis ZA, Herco M, Mathur AM, 2018. Divergent risk factors for cerebellar and intraventricular hemorrhage. J Perinatol 38, 278–284. https://doi.org/10.1038/s41372-017-0010-x

Watterberg, K.L., Gerdes, J.S., Cole, C.H., Aucott, S.W., Thilo, E.H., Mammel, M.C., 2004. Prophylaxis of early adrenal insufficiency to prevent bronchopulmonary dysplasia: a multicenter trial. Pediatrics 114, 1649–57. https://doi.org/10.1542/peds.2004-1159

Watterberg, K.L., Shaffer, M.L., Mishefske, M.J., Leach, C.L., Mammel, M.C., Couser, R.J., Abbasi, S., Cole, C.H., Aucott, S.W., Thilo, E.H., Rozycki, H.J., Lacy, C.B., 2007. Growth and neurodevelopmental outcomes after early low-dose hydrocortisone treatment in extremely low birth weight infants. Pediatrics 120, 40–48. https://doi.org/10.1542/peds.2006-3158

Yaseen, H., Okash, I., Hanif, M., al-Umran, K., al-Faraidy, A., 1999. Early dexamethasone treatment in preterm infants treated with surfactant: a double blind controlled trial. Journal of Tropical Pediatrics 45, 304–6. https://doi.org/10.1093/tropej/45.5.304

Yates, H., Chiocchia, V., Linsell, L., Orsi, N., Juszczak, E., Johnson, K., 2019. Very low-dose dexamethasone to facilitate extubation of preterm babies at risk of bronchopulmonary dysplasia: the MINIDEX feasibility RCT. Efficacy and Mechanism Evaluation 6. https://doi.org/10.3310/eme06080

Yeh, T.F., Lin, Y.J., Hsieh, W.S., Lin, H.C., Lin, C.H., Chen, J.Y., 1997. Early postnatal dexamethasone therapy for the prevention of chronic lung disease in preterm infants with respiratory distress syndrome: a multicenter clinical trial. Pediatrics 100. https://doi.org/10.1542/peds.100.4.e3

Yeh, T.F., Lin, Y.J., Huang, C.C., Chen, Y.J., Lin, C.H., Lin, H.C., 1998. Early dexamethasone therapy in preterm infants: a follow-up study. Pediatrics 101. https://doi.org/10.1542/peds.101.5.e7

Yeh, T.F., Torre, J.A., Rastogi, A., Anyebuno, M.A., Pildes, R.S., 1990. Early postnatal dexamethasone therapy in premature infants with severe respiratory distress syndrome: a double-blind, controlled study. Journal of Pediatrics 117, 273–82. https://doi.org/10.1016/s0022-3476(05)80547-5
